# Does gut microbial richness lost with industrialisation matter for infection? A two-direction test in Gambian toddlers

**DOI:** 10.64898/2026.10.06.26364910

**Authors:** Ogochukwu N. Ofordile

**Affiliations:** MRC Unit The Gambia at the London School of Hygiene & Tropical Medicine, Banjul, The Gambia; Stercora Biosciences Inc., Delaware, USA

**Keywords:** gut microbiome, richness, functional redundancy, insurance hypothesis, industrialisation, childhood infection

## Abstract

**Background and objectives:** Industrialisation is associated with lower gut microbial richness. The ecological insurance hypothesis proposes that richer communities buffer disturbance via overlapping functions, possibly affecting infection susceptibility. We state predictions, audit the evidence, and test the richness-infection relationship in Gambian children.

**Methodology:** We analysed 452 children aged 6–35 months with Day 85 stool 16S profiles and dated infections over 113 days. Diversity used Hill numbers (q = 0, 1, 2); redundancy was inferred from reference-genome gene content. We examined associations with illness during follow-up and infections after Day 85, adjusting for earlier illness.

**Results:** Richness was lower in children with any infection (93.9 vs 103.1; d = 0.36; adjusted odds ratio 0.989 per species, p = 0.019); abundance-weighted diversity was not. Most illness preceded sampling (295 of 315); the difference lay in common and intermediate-prevalence species. Richness was lowest around recent illness. Higher richness was associated with fewer later infections (rate ratio 0.69 per SD, 95% CI 0.51–0.93; q = 0.11). Redundancy correlated with richness at q = 0 (ρ = 0.93); at q = 1–2 estimates indicated lower infection rates but none survived correction (smallest FDR q = 0.14).

**Conclusions and implications:** Richness and infection were associated in both directions: children who had been ill had lower richness, and greater Day 85 diversity preceded fewer infections. Redundancy was directionally aligned with reduced infection at higher diversity orders, but FDR control and collinearity limit inference. Cohorts sampled before illness, with shotgun data, are needed.

**Lay summary:** Gambian toddlers who had been ill had fewer kinds of gut bacteria than those who stayed well. Diversity was lowest around recent illness. Children with more diverse gut communities also experienced fewer infections afterwards. Functional redundancy generally showed the same direction of association, but its contribution beyond richness remains uncertain.

## Background and objectives

Gut microbial communities differ between industrialised and non-industrialised populations. Non-industrialised guts generally carry more taxa and are commonly enriched for *Prevotella*, whereas industrialised guts are less rich and more often *Bacteroides*-dominated [1,2]. The shift can occur rapidly: among 19 Karen individuals sampled before and after migration to the United States, migration was associated with immediate loss of gut microbial diversity and function, with US-associated strains and functions displacing native ones [3]. From an evolutionary-mismatch perspective, such changes matter if they remove ecological functions that contribute to host responses to environmental disturbance.

A specific version of this idea links three properties (Fig. 1a). Richness may supply functional redundancy, with several taxa able to perform overlapping functions; redundancy may confer resilience [4]; and resilience may reduce susceptibility to infection. Across ecosystems, functional redundancy has a positive mean relationship with stability and resilience, although the contribution of species richness itself varies and the underlying mechanisms remain incompletely resolved [5]. In the gut, low diversity and domination predict bacteraemia and mortality after transplantation [6–8], higher recipient redundancy has been associated with resistance to donor engraftment [9], and non-industrialised guts contain richer sets of short-chain fatty acid-producing organisms [10]. The full ecological chain has not been directly tested in a general population of young children.

**Figure 1.**
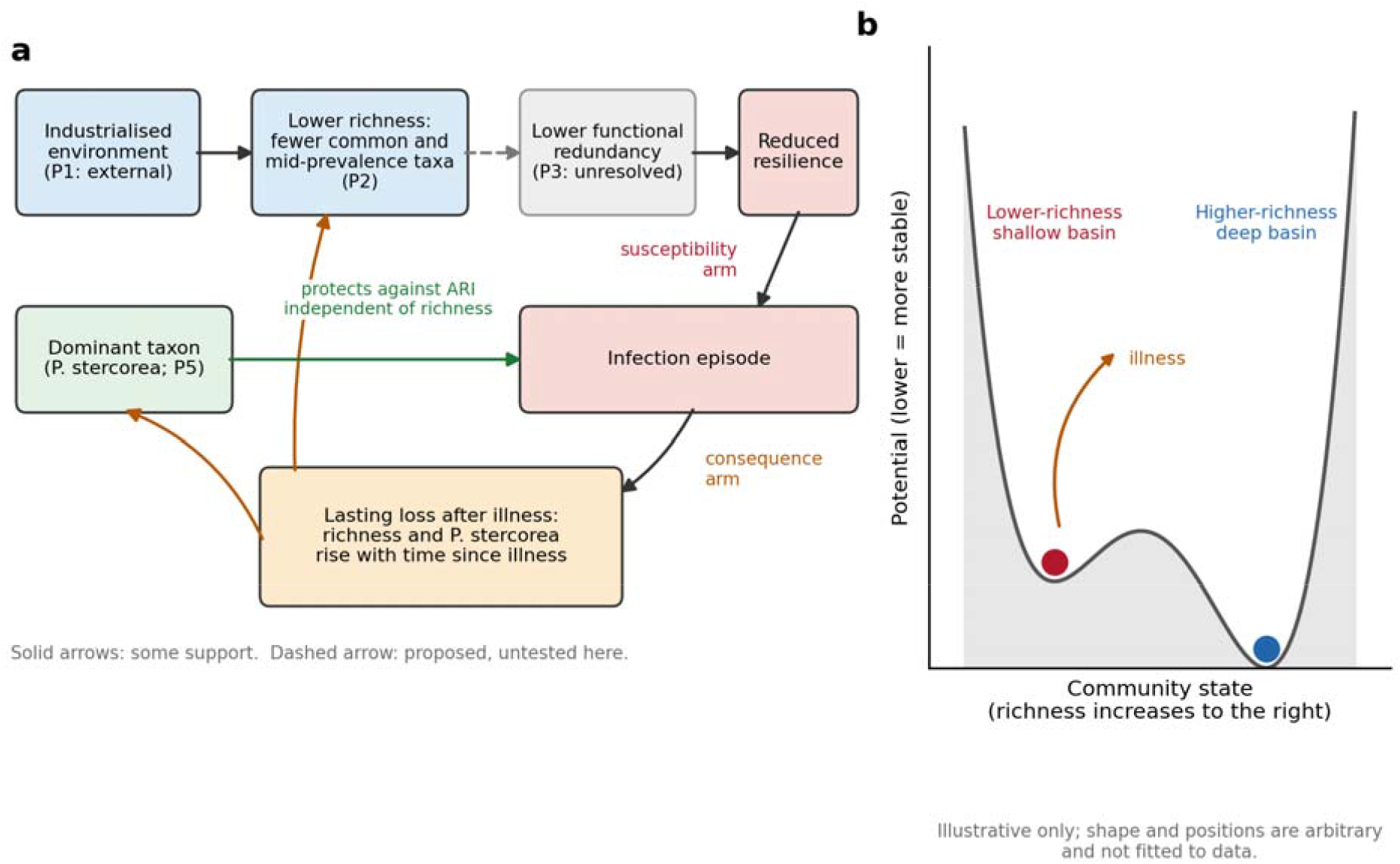
Framework and conceptual landscape. (a) The proposed ecological chain and five predictions (Table 1). The industrialisation-to-richness link is based on previous population studies; the present study tests the richness-infection component and examines redundancy and a taxon-level contrast. The consequence and susceptibility arms address possible bidirectional feedback. (b) Illustrative stability landscape showing a lower-richness community occupying a shallower basin and being more readily displaced by disturbance. Shape and position are conceptual and not fitted to the data.

Richness and abundance-weighted diversity are not interchangeable. Richness (Hill number q = 0) counts every taxon equally; Shannon diversity (q = 1) and inverse Simpson diversity (q = 2) progressively weight abundant taxa more strongly [11]. If the number of taxa itself contributes to ecological insurance, its association with infection should be clearest at q = 0 and weaken as abundance increasingly determines diversity.

**Table 1.**
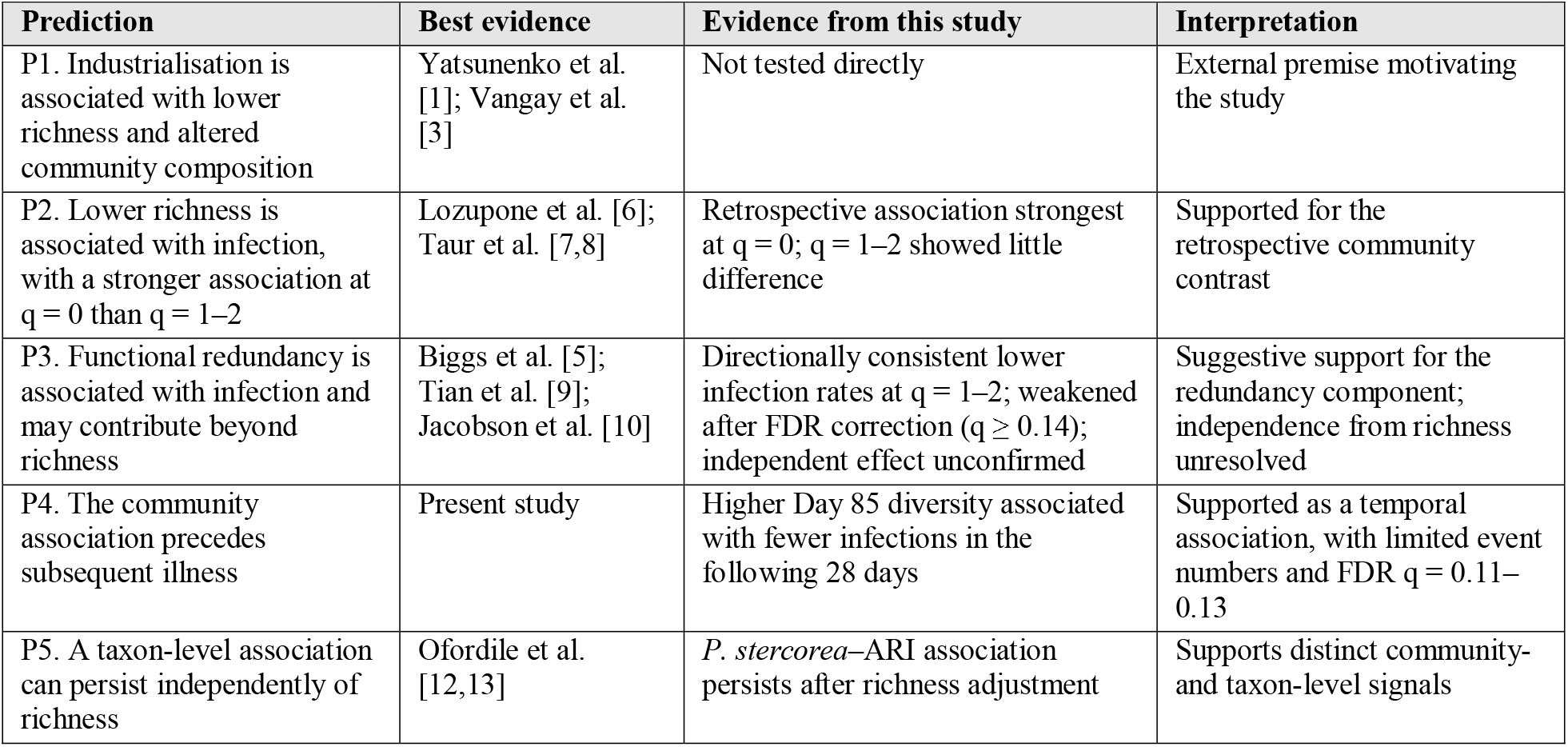
Predictions and evidence audit.

There is also an epidemiological problem. Infection can perturb the gut microbiome, so an association between low richness and infection does not establish direction. Low richness could increase susceptibility, infection could reduce richness, or both could operate in a feedback loop.

We therefore treat the insurance hypothesis as a set of testable predictions rather than a single causal claim. Industrialisation motivates the question, but IHAT-GUT does not compare industrialised and non-industrialised populations. It tests the richness-infection component in a non-industrialised population with dated infection records. We therefore examine two directions: a consequence arm, asking whether previous illness is associated with lower richness and subsequent increase in richness with time since illness; and a susceptibility arm, asking whether diversity before a short post-sampling window is associated with subsequent infection (Fig. 1a). Conceptually, a lower-richness community may occupy a shallower stability basin and be displaced more readily by disturbance (Fig. 1b). We also test whether inferred functional redundancy explains infection associations beyond richness and examine *Prevotella stercorea* as a taxon-level contrast.

## Methodology

### Study population

IHAT-GUT (NCT02941081) was a double-blind, placebo-controlled trial of an iron supplement in 633 children aged 6– 35 months with mild to moderate iron-deficiency anaemia in the Upper River Region of The Gambia during 2017–2018 [14,15]. The Gambia Government/MRC Joint Ethics Committee approved the study (SCC1489), and parents or guardians provided written informed consent, including for subsequent microbiome analyses. Stool was collected on Days 1 (n = 516), 15 (n = 408) and 85 (n = 452) into OMNIgene·GUT tubes. Sample size differences across days reflect attrition and missing sample collection at earlier study days. Severe malnutrition was an exclusion criterion, and samples collected after antibiotic treatment were removed. Adverse events were recorded at five clinics with dates; infection events were classified as acute respiratory infection (ARI), diarrhea or fever.

We analysed Day 85 because illness groups were separated by this point and most recorded illness had already occurred. The analytic set comprised 452 children with a Day 85 sample and illness classification: 137 never ill and 315 with at least one infection. Each event had a Study Day, defined as days since enrolment, with the Day 85 sample treated as study day 85 for all children. Follow-up ended at day 113, establishing a maximum prospective post-sampling window of 28 days.

### Microbiome data

The V1–V2 region of the 16S rRNA gene was sequenced on Illumina MiSeq. Reads were processed with mothur, normalised to 20,000 reads per sample, clustered by minimum entropy decomposition and assigned to species using ARB against a curated SILVA 123 database [12]. The analysis table contained 488 species-level columns of integer counts. The merged species-count and host-metadata table and the event-level adverse-event file are available from Mendeley Data (doi:10.17632/fr5p4cgc7j.2). As a pipeline check, Day 85 richness was also re-derived from forward (R1) reads (vsearch, UNOISE denoising, 99% identity mapping, no taxonomy; analytic rarefaction to 3,000–7,000 reads; Table S12); the ASV table is available from Mendeley Data (doi:10.17632/rb4wd2z7sp.1).

### Diversity and functional redundancy

For each child we calculated Hill numbers of order q = 0, 1 and 2. Richness (q = 0) was the number of species-level taxa with non-zero counts, q = 1 was exponential Shannon diversity and q = 2 was inverse Simpson diversity.

Functional redundancy was calculated as the ratio of the naive to similarity-sensitive Hill number [16], using similarity derived from KEGG Orthology profiles of reference genomes in the PICRUSt2 bacterial reference database [17,18]. The reference contained 26,868 GTDB-based genomes. Each study species was assigned the mean profile of its matched reference genomes, with species-level matches for 259 species, genus-level matches for 220 and no match for 9.

Similarity was measured using the weighted Jaccard index. The similarity matrix covered 479 of 488 study species and approximately 100% of counts per sample. Same-genus pairs were more similar than different-genus pairs (mean weighted Jaccard 0.80 vs 0.38; Fig. 4a,b; Table S10).

## Statistical analysis

Richness and illness were compared using Welch tests and Cohen’s d. Logistic regression was adjusted for age, sex, height-for-age Z-score, trial arm, season, location and assigned counts per sample. Whole-follow-up episode counts were analysed using negative binomial regression (no child had more than five episodes).

Redundancy was added to richness at the corresponding diversity order and then examined with log-transformed *P. stercorea* counts. Benjamini-Hochberg correction was applied across the four outcomes.

The post-sampling analysis included all 452 children and counted infections after study day 85 using quasi-Poisson regression. Models adjusted for age, sex, height-for-age Z-score, trial arm, season, location, assigned counts, infections up to day 85 and any episode within 14 days before the Day 85 sample. Effects were expressed as rate ratios per SD of diversity.

A stricter descriptive comparison contrasted the 20 children whose first infection occurred after day 85 with the 137 children who were never ill. Species were divided by prevalence into common (≥50% of samples; 71 species), intermediate (10–50%; 153 species) and rare (<10%; 264 species). Recovery was examined according to time since the last infection episode ended.

The post-sampling, prevalence-subset and recovery analyses are exploratory. Analyses used R 4.6.1.

## Results

### Most illness preceded the Day 85 sample

Among the 452 children, 137 were never ill and 315 had at least one infection. Of the 315 children with infection, 295 had their first recorded infection on or before study day 85 and only 20 had their first infection afterwards (Fig. 2a–c; Table S1). The main Day 85 comparison therefore primarily contrasts children with previous infection against those without recorded infection.

**Figure 2.**
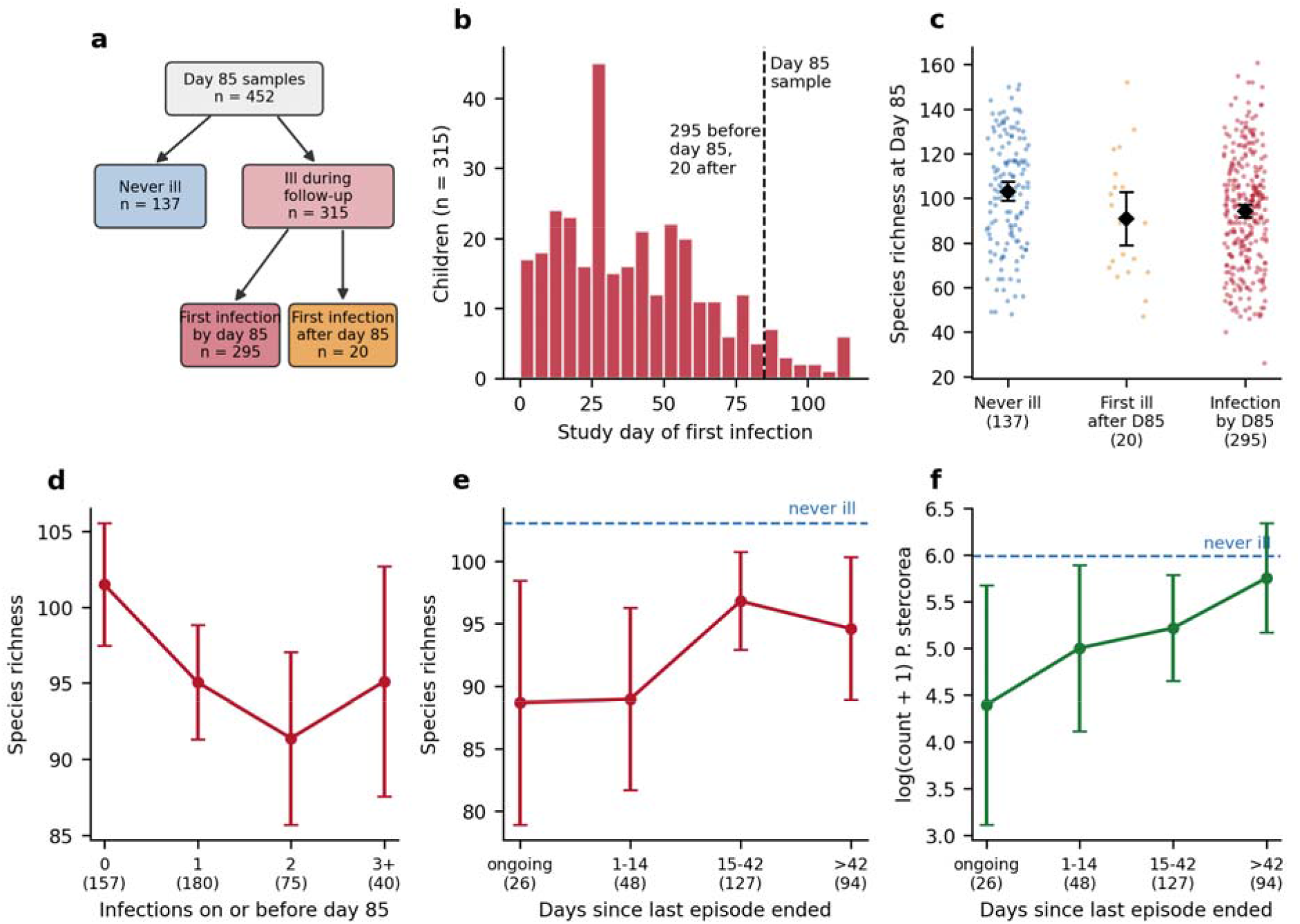
Cohort, timing and the consequence arm. (a) Flow of the 452 children. (b) Study day of first infection in the 315 children with infection; dashed line, the Day 85 sample. (c) Species richness by timing group, means with 95% CI. (d) Richness by number of infections on or before day 85. (e,f) Richness and log(count + 1) *P. stercorea* by days since the last episode ended; dashed lines, never-ill means. Panels d–f are exploratory.

### Richness, not abundance-weighted diversity, differs

Day 85 richness was lower in children with infection than in never-ill children (93.9 ± 25.5 vs 103.1 ± 25.3 species; Cohen’s d = 0.36, 95% CI 0.16–0.56; Welch p = 0.0005). Abundance-weighted measures showed no corresponding difference: exponential Shannon diversity was 10.7 vs 10.5 (d = −0.05, 95% CI −0.25 to 0.15) and inverse Simpson diversity was 5.5 vs 5.1 (d = −0.12, 95% CI −0.32 to 0.08) (Fig. 3a; Table S2).

**Figure 3.**
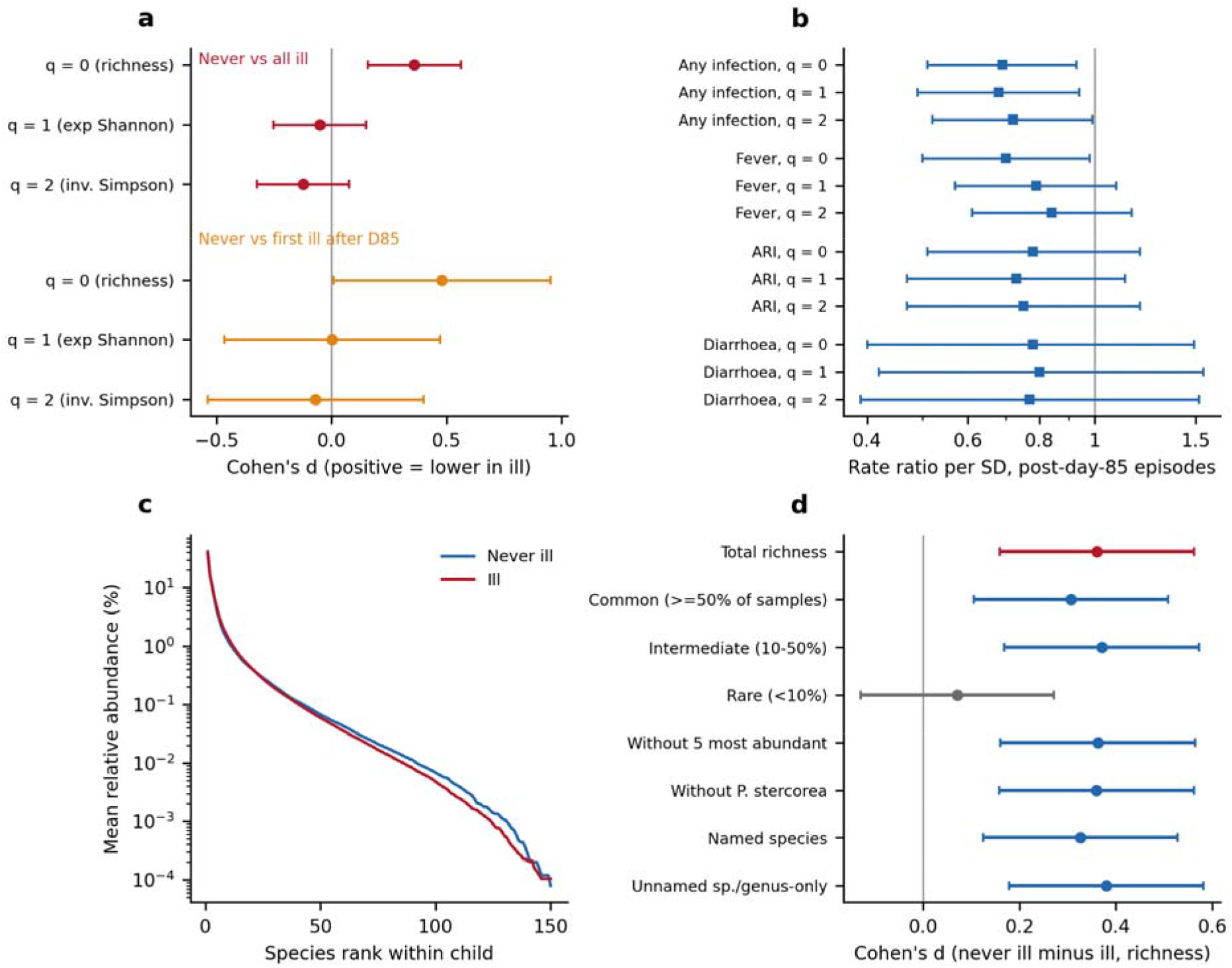
Richness, evenness and prospective infection. (a) Cohen’s d for the difference between never-ill and infection groups at q = 0, 1 and 2; positive values indicate lower diversity in children with infection. The 20 children first ill after day 85 are shown separately against the 137 never-ill children. (b) Post-sampling rate ratios per SD of diversity for post-day-85 infection episodes in all 452 children, adjusted for earlier illness. (c) Mean relative abundance by species rank within child. (d) Cohen’s d for richness in common, intermediate and rare species, after exclusion of dominant taxa and for named and unnamed species. Bars show 95% CI.

The association between richness and infection remained after adjustment (odds ratio 0.989 per additional species, 95% CI 0.979–0.998, p = 0.019, n = 442), rarefaction to 10,000 counts (p = 0.043) and removal of species present in fewer than 5% of samples (p = 0.024) (Fig. S1). Adjusted richness differences at Days 1 and 15 were smaller and included zero (Table S3). Whole-follow-up episode counts gave weaker associations, with incidence rate ratios of 0.992–0.998 per species (Table S4). An independent re-derivation from the R1 reads using a separate ASV pipeline reproduced the association (rarefied ASV richness, d = 0.39; adjusted odds ratio 0.75 per SD, 95% CI 0.58–0.97; post-sampling rate ratio 0.68 per SD, 0.50–0.94; Table S12).

### The difference lies in common and intermediate-prevalence species

The richness difference was carried by common species (d = 0.31, 95% CI 0.11–0.51) and intermediate-prevalence species (d = 0.37, 95% CI 0.17–0.57), whereas the difference for rare species was small and included zero (d = 0.07, 95% CI −0.13 to 0.27) (Fig. 3d; Table S7).

The difference remained after removal of the five most abundant species or *P. stercorea* (d = 0.36 in each analysis) and was similar for named species (d = 0.33) and unnamed species (d = 0.38). Rank-abundance curves differed mainly through a lower mid-rank tail in children with infection (Fig. 3c). The signal therefore does not depend on singleton assignments or a small number of dominant taxa.

### Consequence arm: richness is lowest around recent illness

Richness did not show a graded relationship with the number of previous infection episodes. Mean richness was 101.5, 95.0, 91.4 and 95.1 species in children with 0, 1, 2 and ≥3 episodes, respectively. The adjusted change was −1.1 species per additional episode (95% CI −3.3 to 1.2, p = 0.35; Table S8).

Among children ill by Day 85, richness exhibited a temporal recovery gradient rather than a cumulative burden response. Mean richness was lowest during active or recent illness within 14 days (88.7–89.0 species) and higher at 15–42 days (96.8 species) and beyond 42 days (94.6 species), relative to 103.1 in never-ill controls (Fig. 2e). Log-transformed *P. stercorea* abundance mirrored this recovery pattern (4.4, 5.0, 5.2, and 5.8 across timing groups vs. 6.0 in controls) (Fig. 2f).

The temporal gradient is consistent with a loss of richness around illness followed by subsequent recovery. The pattern does not show a graded relationship with cumulative illness burden.

### Susceptibility arm: higher Day 85 diversity is associated with fewer subsequent infections

Across all 452 children, higher Day 85 diversity was inversely associated with subsequent infection episodes during the 28-day follow-up after adjustment for earlier illness. Rate ratios per standard deviation of diversity were 0.69 (95% CI 0.51–0.93, p = 0.015) for species richness (q = 0), 0.68 (95% CI 0.49–0.94, p = 0.019) for exponential Shannon diversity (q = 1), and 0.72 (95% CI 0.52–0.99, p = 0.044) for inverse Simpson diversity (q = 2). Fever gave a rate ratio of 0.70 (95% CI 0.50–0.98, p = 0.038) for richness, while ARI and diarrhoea pointed in the same direction with wider intervals (Fig. 3b; Table S6).

After Benjamini-Hochberg correction across the 12 diversity tests, q values for any infection were 0.114 (q = 0), 0.114 (q = 1), and 0.132 (q = 2). Thus the prospective association did not survive FDR correction. Importantly, the direction was consistent across all three diversity orders.

In the stricter descriptive comparison, the 20 children whose first infection occurred after day 85 had mean richness of 90.8 species compared with 103.1 in the 137 never-ill children (d = 0.48, 95% CI 0.01–0.95; Welch p = 0.075).

### Functional redundancy shows a consistent directional association

Inferred functional redundancy was strongly correlated with richness at q = 0 (Spearman ρ = 0.93), less correlated at q = 1 (ρ = 0.63) and more separable at q = 2 (ρ = 0.33) (Fig. 4c–e).

**Figure 4.**
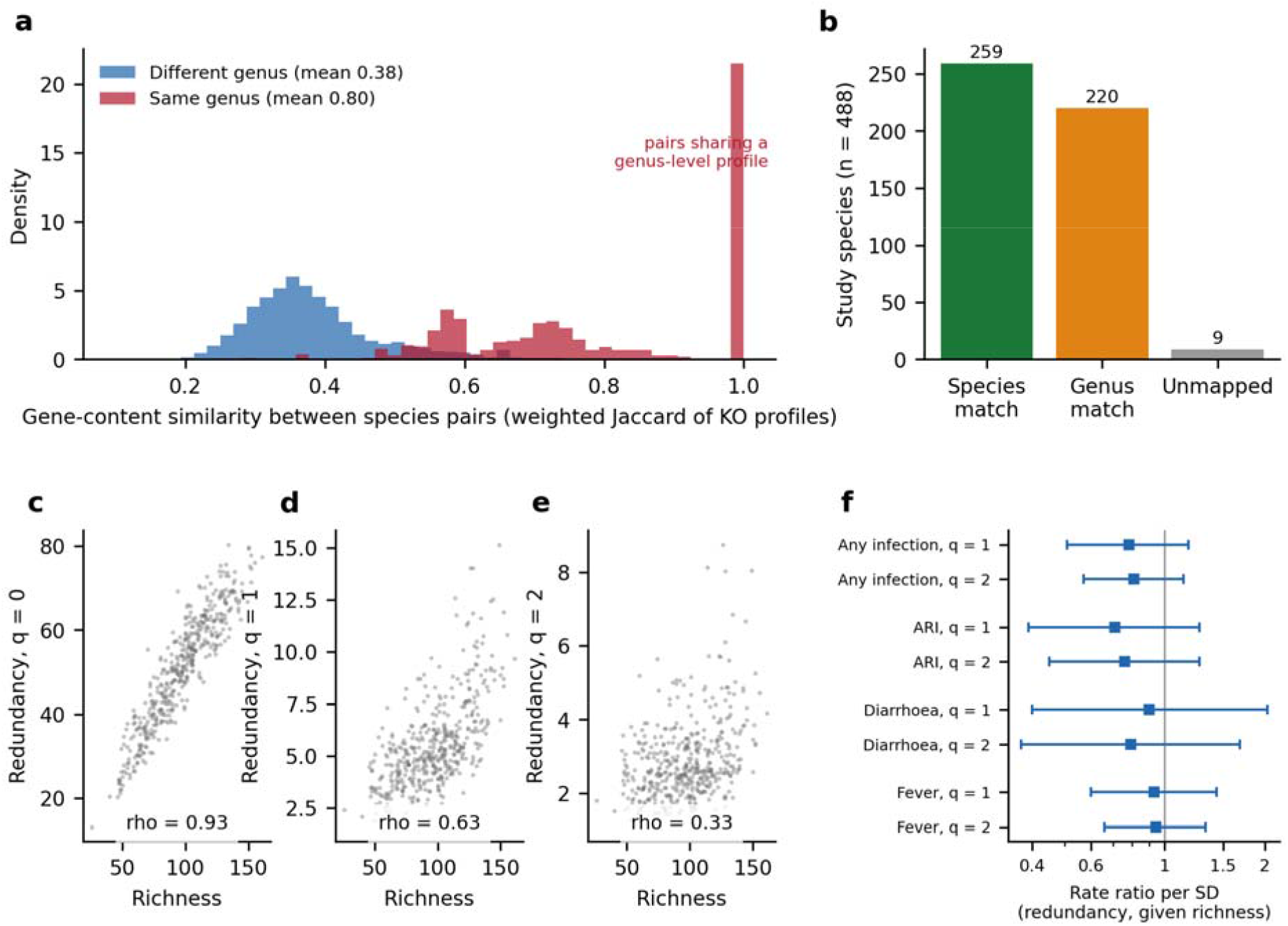
Similarity matrix and redundancy analysis. (a) Pairwise gene-content similarity between study species, measured by weighted Jaccard similarity of KEGG Orthology profiles, within and between genera. (b) Study species matched to reference genomes at species or genus level, or unmapped. (c–e) Inferred redundancy against richness at q = 0, 1 and 2, with Spearman ρ. (f) Post-sampling rate ratios per SD of redundancy conditional on richness for subsequent infection episodes, with 95% CI.

At q = 0, redundancy added little information beyond richness because of severe collinearity. At q = 1 and q = 2, estimates generally pointed in the predicted direction.

For ARI at q = 2, the rate ratio was 0.48 (95% CI 0.23–1.02, p = 0.054; Table S5). For all illness it was 0.60 (95% CI 0.35–1.05, p = 0.070). These estimates are directionally consistent with lower infection rates at greater redundancy, but their confidence intervals include 1 and multi-testing correction yielded FDR q-values ≥ 0.14 in the negative binomial models (≥ 0.12 in quasi-Poisson refits; Table S5), leaving an effect independent of richness unestablished. Restricting the similarity matrix to the 259 species with species-level genome matches produced estimates near 1.

In the post-sampling analysis, redundancy conditional on richness gave rate ratios of 0.78 (95% CI 0.51–1.18, p = 0.24) at q = 1 and 0.81 (95% CI 0.57–1.14, p = 0.23) at q = 2 for any subsequent infection.

Adding *P. stercorea* attenuated the q = 2 redundancy estimate for ARI from 0.48 to 0.68 (p = 0.34). This indicates that part of the apparent redundancy signal may be related to taxon-level structure, although the analysis does not establish mediation.

### *P. stercorea* provides a distinct taxon-level signal

The community-level richness association is not reducible to *P. stercorea*: removing *P. stercorea* left the richness effect unchanged (d = 0.36). Conversely, earlier analyses of this cohort found that *P. stercorea* was associated with lower ARI frequency after adjustment for richness [12,13]. In earlier analyses, the *P. stercorea* association remained essentially unchanged after richness adjustment, with IRR 0.942 (95% CI 0.907–0.979), compared with 0.946 before adjustment [13]. In a joint model with *P. copri, P. stercorea* retained its association whereas *P. copri* did not [13] (Table S9).

These results distinguish two levels of ecological signal: a community-level association involving richness and a taxon-level association involving *P. stercorea*. The present study does not establish that *P. stercorea* causes protection, but neither can its association be explained simply by community richness.

## Discussion

The central finding is that gut community richness and infection show a temporal pattern compatible with a two-direction relationship.

At Day 85, children who had experienced infection had fewer taxa than children who remained infection-free: 93.9 versus 103.1 species, with a moderate standardized difference (d = 0.36). The difference was specific to richness rather than Shannon or inverse Simpson diversity and was concentrated in common and intermediate-prevalence taxa. It persisted after adjustment, rarefaction, prevalence filtering and removal of dominant taxa.

The timing of illness adds an important dimension. Richness was lowest during or shortly after illness and was higher at longer intervals after the last episode. The pattern is consistent with infectious disturbance followed by partial recovery. It also provides a plausible explanation for why the principal Day 85 contrast is difficult to interpret causally: much of the microbiome difference may be a consequence of earlier illness.

The susceptibility arm addresses the reverse direction. Higher Day 85 diversity was associated with fewer infections during the subsequent 28 days, even after adjustment for earlier illness. The association was consistent across richness, Shannon and inverse Simpson diversity. It did not survive the Benjamini-Hochberg threshold across 12 tests (q = 0.11– 0.13), but the temporal ordering and concordant direction across diversity measures make it relevant to the overall pattern.

Together, the two arms support a model in which infectious disturbance and gut community state may influence one another. Infection can be followed by a reduction in richness, while a more diverse community may subsequently experience fewer infection episodes.

### What the findings say about ecological insurance

The ecological insurance hypothesis provides a useful framework, but the data distinguish several components of it.

The retrospective association has the expected richness-specific form. The difference is apparent at q = 0 but not at q = 1 or q = 2. This is consistent with a role for the number of taxa rather than simply the abundance distribution of dominant organisms.

The prospective association is broader. Richness, Shannon and inverse Simpson diversity all show similar rate ratios for subsequent infection. Thus the forward association is not uniquely attributable to richness. A broader community state may be involved.

Functional redundancy provides a further, and more nuanced, signal. Several redundancy estimates were directionally consistent with lower infection rates, although FDR-corrected q-values exceeded 0.14. This pattern leans toward the proposed ecological mechanism rather than providing evidence against it. However, redundancy was highly correlated with richness, particularly at q = 0, and adjustment for richness did not produce a clearly independent association. The data therefore support a possible redundancy contribution while leaving its independence from richness unresolved.

### Functional redundancy: suggestive rather than absent

The q = 2 ARI model illustrates this nuance: while pointing to a substantial inverse association (RR = 0.48, 95% CI 0.23– 1.02, p = 0.054), multi-testing adjustments (q ≥ 0.14) and wide confidence intervals prevent ruling out chance variation. Rather than demonstrating an absence of effect, these findings show a qualitative directional concordance across outcomes, while statistical precision and collinearity prevent isolating an independent contribution of redundancy from richness.

Functional redundancy was inferred from 16S taxonomic assignments rather than measured directly, and 220 mapped species were represented by genus-level rather than species-level reference profiles. Species-level-only analyses produced estimates closer to 1. Overall gene-content similarity may also be an imperfect proxy for redundancy in specific ecological functions. Cross-feeding, niche occupation and colonisation resistance can depend on interactions among organisms whose overall gene repertoires are not interchangeable.

The next test should therefore combine prospective shotgun metagenomics with function-specific measures of redundancy and sampling before, during and after infection.

### Community richness and *P. stercorea* are complementary signals

The *P. stercorea* result is important for interpreting the richness association.

Removing *P. stercorea* from community profiles leaves the overall richness difference unchanged (d = 0.36). Conversely, the protective association between *P. stercorea* and acute respiratory infection persists following adjustment for total richness (IRR = 0.942, 95% CI 0.907–0.979). These findings are consistent with distinct community-level and taxon-level signals.

### Industrialisation and the evolutionary question

The industrialisation question arises from a population-level pattern: industrialised populations tend to have less rich gut communities and different dominant microbial ecologies [1–3]. Vangay et al. provided particularly direct evidence by following individuals before and after migration to the United States, showing rapid loss of diversity and displacement of native strains and functions [3]. The Gambian microbiome provides an important contrast, with a developing, *Prevotella*-rich trophic network that is poorly represented in Western populations [2].

IHAT-GUT does not test whether industrialisation causes infection. It tests a more specific component of the proposed evolutionary argument: whether community richness is related to infection and to the trajectory of the gut ecosystem around infectious disturbance in a non-industrialised population.

If richness falls after infection and higher diversity is associated with fewer subsequent infections, then a population-level reduction in richness could plausibly alter the ecological context in which infectious disturbances occur. Establishing that link requires longitudinal cohorts sampled before illness, during infection and throughout recovery, ideally spanning populations with different degrees of industrialisation.

### Spaceflight as an independent perturbation

Spaceflight provides a useful example of microbiome perturbation without serving as validation of the present hypothesis. In the International Space Station study, the intestinal microbiota changed during long-duration missions, with convergence between astronauts driven largely by changes in several taxa [19]. The NASA Twins Study likewise found substantial microbiome changes during spaceflight without a simple loss of overall diversity [20] (Table S11).

These observations show that ecological perturbation can alter community composition without necessarily reducing richness. They therefore support treating richness, composition and functional capacity as distinct ecological dimensions.

### Strengths and limitations

The principal strength is the combination of dated infection records with microbiome sampling in young children from a non-industrialised setting. The Day 85 sample provides a division between the large body of previous illness and a defined 28-day prospective window. The richness association is also robust to sequencing-depth, prevalence and dominant-taxon analyses.

The principal limitation is the number and timing of prospective events. Only 20 children had their first infection after Day 85, and the post-sampling window was at most 28 days. The recovery analysis is observational rather than a within-child experimental demonstration of recovery. The cohort was originally assembled for an iron trial, and residual confounding cannot be excluded.

A second limitation is measurement. Richness is based on species-level 16S assignments, while functional redundancy is inferred from reference genomes. The infection outcome is based on clinically recorded events and combines several syndromes.

## Conclusions and implications

In Gambian toddlers, gut microbial richness and infection show associations in both directions. Children who had experienced infection had lower Day 85 richness, particularly among common and intermediate-prevalence taxa. Richness was lowest around recent illness and was higher at longer intervals after the last episode. Higher Day 85 diversity was also associated with fewer infection episodes during the following 28 days.

The pattern is consistent with a feedback relationship in which infection can perturb community richness while community state may influence subsequent infection. The retrospective association is particularly specific to richness, whereas the prospective association extends across diversity orders.

Functional redundancy showed directionally consistent associations with lower infection rates at higher diversity orders (q = 1, 2). However, strict FDR control (q ≥ 0.14) and collinearity with taxon richness limit inferential confidence, leaving an independent redundancy effect unestablished in this cohort.

The findings also distinguish community and taxon levels of association. *P. stercorea* remains associated with lower ARI frequency after richness adjustment, while the richness association remains after removal of *P. stercorea*. These observations indicate that community richness and taxon-specific associations capture different aspects of the microbiome-infection relationship.

The broader evolutionary question is whether the richness loss associated with industrialisation carries consequences for infectious resilience. This study does not test industrialisation directly. It identifies the ecological relationship that a direct test should now address. Prospective sampling before illness across populations with different degrees of industrialisation, combined with shotgun metagenomics and function-specific measures of redundancy, would provide that test.

## Data Availability

All data used are publicly available. Analysis input files are on Mendeley Data (doi:10.17632/fr5p4cgc7j.2 and doi:10.17632/rb4wd2z7sp.1); raw 16S rRNA gene reads are in the European Nucleotide Archive (ERP110905). Analysis code is available at https://github.com/ofordile-star/western-microbiome-fragility.

https://doi.org/10.17632/fr5p4cgc7j.2

https://doi.org/10.17632/rb4wd2z7sp.1

https://www.ebi.ac.uk/ena/browser/view/ERP110905

https://github.com/ofordile-star/western-microbiome-fragility

## Declarations

### Ethics

IHAT-GUT (ClinicalTrials.gov NCT02941081) was approved by the Gambia Government/MRC Joint Ethics Committee (SCC1489) and under Clinical Trials Authorisation HP373/347/16/MJK(80). Written informed consent was obtained from parents or guardians.

### Data availability

Analysis input files (merged microbiome–host table, adverse events): Mendeley Data, doi:10.17632/fr5p4cgc7j.2; R1-only ASV table: Mendeley Data, doi:10.17632/rb4wd2z7sp.1. Raw 16S reads: European Nucleotide Archive ERP110905.

### Code Availability

Analysis code and figure generation scripts are publicly available at https://github.com/ofordile-star/western-microbiome-fragility and will be formally archived at Zenodo upon acceptance.

### Related preprint

An earlier, substantially different version of this work, with a different title, framing and set of analyses (including a spaceflight dataset), was posted at Research Square during review at another journal (https://www.researchsquare.com/article/rs-9981104/v1). The present manuscript uses the same IHAT-GUT dataset but reports a different hypothesis, a different set of analyses (a two-direction test including similarity-sensitive redundancy) and revised conclusions.

### Software and reproducibility

Analyses were performed in Python 3.12 (pandas, numpy, scipy, pyreadr and matplotlib) and R 4.6.1 using RStudio 2026.07.1 and Quarto 1.9.38. Weighted Jaccard similarity was calculated from KEGG Orthology profiles using PICRUSt2 reference files pinned to a specific commit and verified by SHA-256 checksums.

Regression models used tidyverse, MASS, broom and lubridate. Hill numbers and the similarity-sensitive redundancy ratio were calculated using custom R functions following Leinster and Cobbold. Welch tests, Cohen’s *d* and Benjamini– Hochberg correction were performed in Python; logistic and negative binomial regression in R. Figures were generated in matplotlib and formatted in Inkscape 1.4.2. Input checksums were recorded, and an independent validation script compared regenerated key values with those reported in the manuscript.

### Author contributions

O.N.O.: Conceptualization, Methodology, Software, Validation, Formal analysis, Investigation, Data curation, Visualization, Writing – original draft, Writing – review & editing, Project administration.

## References

1. Yatsunenko T, Rey FE, Manary MJ, et al. Human gut microbiome viewed across age and geography. Nature. 2012;486:222–227.

2. de Goffau MC, Jallow AT, Sanyang C, et al. Gut microbiomes from Gambian infants reveal the development of a non-industrialised Prevotella-based trophic network. Nat Microbiol. 2022;7:132–144.

3. Vangay P, Johnson AJ, Ward TL, et al. US immigration westernizes the human gut microbiome. Cell. 2018;175:962–972.

4. Yachi S, Loreau M. Biodiversity and ecosystem productivity in a fluctuating environment: the insurance hypothesis. Proc Natl Acad Sci USA. 1999;96:1463–1468.

5. Biggs CR, Yeager LA, Bolser DG, et al. Does functional redundancy affect ecological stability and resilience? A review and meta-analysis. Ecosphere. 2020;11:e03184.

6. Lozupone CA, Stombaugh JI, Gordon JI, Jansson JK, Knight R. Diversity, stability and resilience of the human gut microbiota. Nature. 2012;489:220–230.

7. Taur Y, Jenq RR, Perales MA, et al. The effects of intestinal tract bacterial diversity on mortality following allogeneic hematopoietic stem cell transplantation. Blood. 2014;124:1174–1182.

8. Taur Y, Xavier JB, Lipuma L, et al. Intestinal domination and the risk of bacteremia in patients undergoing allogeneic hematopoietic stem cell transplantation. Clin Infect Dis. 2012;55:905–914.

9. Tian L, Wang XW, Wu AK, et al. Deciphering functional redundancy in the human microbiome. Nat Commun. 2020;11:6217.

10. Jacobson DK, Honap TP, Ozga AT, et al. Analysis of global human gut metagenomes shows that metabolic resilience potential for short-chain fatty acid production is strongly influenced by lifestyle. Sci Rep. 2021;11:1724.

11. Chao A, Chiu C-H, Jost L. Unifying species diversity, phylogenetic diversity, functional diversity, and related similarity and differentiation measures through Hill numbers. Annu Rev Ecol Evol Syst. 2014;45:297–324.

12. Ofordile O, Pereira DIA, Prentice AM, Wagner J. Gut Prevotella stercorea associates with protection against infection in rural African children. Nat Commun. 2025;16:11101.

13. Ofordile O. Prevotella stercorea links gut microbiome ecology to respiratory infection protection through host-context-dependent, richness-independent associations. BMC Med Genomics. 2026. doi:10.1186/s12920-026-02449-9.

14. Mohammed NI, et al. A novel nano-iron supplement versus standard treatment for iron deficiency anaemia in children 6–35 months (IHAT-GUT trial): a double-blind, randomised, placebo-controlled non-inferiority phase II trial in The Gambia. EClinicalMedicine. 2023. doi:10.1016/j.eclinm.2023.101853.

15. Pereira DIA, et al. A novel nano-iron supplement to safely combat iron deficiency and anaemia in young children: the IHAT-GUT double-blind, randomised, placebo-controlled trial protocol. Gates Open Res. 2018. doi:10.12688/gatesopenres.12866.2.

16. Leinster T, Cobbold CA. Measuring diversity: the importance of species similarity. Ecology. 2012;93:477–489.

17. Douglas GM, Maffei VJ, Zaneveld JR, et al. PICRUSt2 for prediction of metagenome functions. Nat Biotechnol. 2020;38:685– 688.

18. Parks DH, Chuvochina M, Rinke C, et al. GTDB: an ongoing census of bacterial and archaeal diversity through a phylogenetically consistent, rank normalized and complete genome-based taxonomy. Nucleic Acids Res. 2022;50:D785–D794.

19. Voorhies AA, Ott CM, Mehta S, et al. Study of the impact of long-duration space missions at the International Space Station on the astronaut microbiome. Sci Rep. 2019;9:9911.

20. Garrett-Bakelman FE, Darshi M, Green SJ, et al. The NASA Twins Study: a multidimensional analysis of a year-long human spaceflight. Science. 2019;364:eaau8650.

