## Supplement for "Does gut microbial richness lost with industrialisation matter for infection? A two-direction test in Gambian toddlers"

### Methods S1. Similarity matrix and definitions

Genome-level KEGG-orthology (KO) annotations and GTDB taxonomy were taken from the PICRUSt2 default bacterial reference (github.com/picrust/picrust2; picrust2/default_files/bacteria) [1,2]. Each of the 488 study species was matched by name to genomes (species-level for 259, genus-level for 220, none for 9); a species' profile is the mean KO presence frequency across its genomes (4,222 genomes used). Similarity between two species is the weighted Jaccard index of their profiles (sum of minima over sum of maxima), with 1 on the diagonal, used as the similarity matrix of the Leinster–Cobbold similarity-sensitive Hill number [3]. Redundancy is the naive Hill number divided by the similarity-sensitive Hill number at the same order. The matrix has 479 species. Build script: 01a_build_similarity.py.

Event definitions: infection, ARI, diarrhoea and fever are flags in the adverse-event file; Study Day is days since enrolment; pre-day-85 events have Study Day ≤ 85; episode duration is end date minus start date plus one day. Software: R 4.6.1 (tidyverse, MASS, broom) and Python 3. Scripts: 01b_DataPrep.R, 02_main_models.R, 03_post_sampling.R, 04_child_level_data.R, 05_additional_tests.R, 09_asv_replication.R (with config.R and fr_helpers.R), and 01a_build_similarity.py.

### Supplementary tables

**Table S1.** Characteristics of the analytic set by timing of first infection.

| | Never ill | First ill after day 85 | Ill by day 85 | Total |
|---|---|---|---|---|
| n | 137 | 20 | 295 | 452 |
| Age at enrolment, months, mean (SD) | 24.0 (8.1) | 22.6 (7.5) | 21.6 (8.1) | 22.4 (8.1) |
| Height-for-age Z-score, mean (SD) | −0.92 (0.96) | −0.79 (0.85) | −0.87 (0.94) | −0.88 (0.94) |
| Treatment arm, % | 67.2 | 80.0 | 67.1 | 67.7 |
| Species richness, mean (SD) | 103.1 (25.3) | 90.8 (27.9) | 94.1 (25.3) | 96.7 (26.0) |

**Table S2.** Richness and evenness-weighted diversity at Day 85 (species level).

| Measure | Never ill (n = 137) | Ill (n = 315) | Cohen's d (95% CI) | Welch p |
|---|---|---|---|---|
| Richness (q = 0) | 103.06 ± 25.28 | 93.90 ± 25.47 | 0.36 (0.16–0.56) | 0.0005 |
| Shannon index | 2.251 ± 0.439 | 2.272 ± 0.452 | −0.05 (nominally higher in ill) | 0.649 |
| Exponential Shannon (q = 1) | 10.46 | 10.71 | −0.05 (−0.25 to 0.15) | 0.617 |
| Inverse Simpson (q = 2) | 5.14 | 5.46 | −0.12 (−0.32 to 0.08) | 0.217 |

Area under the curve for illness: richness 0.605 (0.548–0.662), Shannon 0.521 (0.463–0.578); DeLong p = 0.104 (earlier analysis of this cohort).

**Table S3.** Adjusted richness difference (never ill minus ill) by timepoint; linear model with illness tested after age group (three categories).

| Timepoint | n | Difference in richness (95% CI) | F | p |
|---|---|---|---|---|
| Day 1 | 516 | 3.3 (−1.0 to 7.7) | 2.29 | 0.13 |
| Day 15 | 408 | 3.2 (−1.6 to 8.0) | 1.74 | 0.19 |
| Day 85 | 452 | 5.9 (1.2 to 10.6) | 6.19 | 0.013 |

**Table S4.** Whole-follow-up infection episodes (negative binomial, capped at 5): incidence rate ratio per unit of diversity (95% CI; p).

| Outcome | Richness (q = 0) | q = 1 | q = 2 |
|---|---|---|---|
| ARI | 0.998 (0.993–1.004; 0.559) | 0.984 (0.959–1.008; 0.196) | 0.972 (0.926–1.017; 0.237) |
| Diarrhoea | 0.992 (0.984–1.001; 0.075) | 0.992 (0.952–1.030; 0.680) | 0.994 (0.919–1.066; 0.869) |
| All illness | 0.997 (0.993–1.001; 0.141) | 0.988 (0.970–1.006; 0.202) | 0.978 (0.944–1.012; 0.222) |
| Illness excluding ARI | 0.995 (0.989–1.001; 0.106) | 0.993 (0.966–1.020; 0.632) | 0.986 (0.934–1.037; 0.601) |

**Table S5.** Redundancy and whole-follow-up infection episodes, adjusted for richness at the same order: rate ratio per unit of the redundancy ratio (95% CI; p).

| Outcome | q = 0 | q = 1 | q = 2 |
|---|---|---|---|
| ARI (control) | 1.01 (0.98–1.03; 0.54) | 0.78 (0.56–1.09; 0.15) | 0.48 (0.23–1.02; 0.054) |
| Diarrhoea | 0.99 (0.95–1.02; 0.46) | 0.75 (0.45–1.27; 0.27) | 0.55 (0.17–1.79; 0.31) |
| All illness | 1.00 (0.98–1.02; 0.93) | 0.84 (0.66–1.08; 0.18) | 0.60 (0.35–1.05; 0.07) |
| Illness excluding ARI | 0.99 (0.96–1.02; 0.40) | 0.93 (0.64–1.35; 0.69) | 0.79 (0.35–1.82; 0.57) |

No negative binomial M2 or M3 result has a corrected q below 0.14. Quasi-Poisson refits give essentially the same estimates (smallest corrected q = 0.12; none below 0.05). A matrix restricted to the 259 species with species-level genome matches gives estimates near 1 (q = 1: 0.97–1.10; q = 2: 0.95–1.06).

**Table S6.** Post-sampling analysis (exploratory): post-day-85 infection episodes in all 452 children (quasi-Poisson), adjusted for age, sex, height-for-age, arm, season, location, assigned counts, infections to day 85 and any episode within 14 days before the sample. Rate ratio per SD (95% CI; p).

| Outcome | Exposure | Rate ratio | 95% CI | p |
|---|---|---|---|---|
| Infection | richness q = 0 | 0.69 | 0.51–0.93 | 0.015 |
| Infection | q = 1 | 0.68 | 0.49–0.94 | 0.019 |
| Infection | q = 2 | 0.72 | 0.52–0.99 | 0.044 |
| Infection | redundancy q = 1, given richness | 0.78 | 0.51–1.18 | 0.242 |
| Infection | redundancy q = 2, given richness | 0.81 | 0.57–1.14 | 0.227 |
| ARI | richness q = 0 | 0.78 | 0.51–1.20 | 0.262 |
| ARI | q = 1 | 0.73 | 0.47–1.13 | 0.160 |
| ARI | q = 2 | 0.75 | 0.47–1.20 | 0.234 |
| ARI | redundancy q = 1, given richness | 0.71 | 0.39–1.27 | 0.247 |
| ARI | redundancy q = 2, given richness | 0.76 | 0.45–1.27 | 0.287 |
| Diarrhoea | richness q = 0 | 0.78 | 0.40–1.49 | 0.446 |
| Diarrhoea | q = 1 | 0.80 | 0.42–1.55 | 0.515 |
| Diarrhoea | q = 2 | 0.77 | 0.39–1.52 | 0.452 |
| Diarrhoea | redundancy q = 1, given richness | 0.90 | 0.40–2.04 | 0.801 |

| Outcome | Exposure | Rate ratio | 95% CI | p |
|---|---|---|---|---|
| Diarrhoea | redundancy q = 2, given richness | 0.79 | 0.37–1.68 | 0.544 |
| Fever | richness q = 0 | 0.70 | 0.50–0.98 | 0.038 |
| Fever | q = 1 | 0.79 | 0.57–1.09 | 0.150 |
| Fever | q = 2 | 0.84 | 0.61–1.16 | 0.285 |
| Fever | redundancy q = 1, given richness | 0.93 | 0.60–1.43 | 0.734 |
| Fever | redundancy q = 2, given richness | 0.94 | 0.66–1.33 | 0.712 |

Episodes: any infection 62 in 53 children; ARI 28 in 25; diarrhoea 15 in 14; fever 47 in 39. Benjamini–Hochberg q across 12 diversity tests: any infection 0.114 (q = 0), 0.114 (q = 1), 0.132 (q = 2); fever (q = 0) 0.132.

**Table S7.** Anatomy of the richness difference at Day 85 (exploratory), never ill vs ill (137 vs 315).

| Species subset | Never ill | Ill | Cohen's d (95% CI) | p (BH q) |
|---|---|---|---|---|
| Total richness | 103.1 | 93.9 | 0.36 (0.16 to 0.56) | <0.001 (<0.001) |
| Common species (>=50% of samples; n=71) | 54.0 | 50.5 | 0.31 (0.11 to 0.51) | 0.002 (0.002) |
| Intermediate (10-50%; n=153) | 41.4 | 36.0 | 0.37 (0.17 to 0.57) | <0.001 (<0.001) |
| Rare (<10%; n=264) | 7.7 | 7.4 | 0.07 (-0.13 to 0.27) | 0.486 (0.486) |
| Richness without 5 most abundant species | 98.5 | 89.3 | 0.36 (0.16 to 0.56) | <0.001 (<0.001) |
| Richness without P. stercorea | 102.2 | 93.0 | 0.36 (0.16 to 0.56) | <0.001 (<0.001) |
| Named species (n=357) | 70.9 | 65.4 | 0.33 (0.12 to 0.53) | 0.001 (0.002) |
| Unnamed sp./genus-only (n=131) | 32.2 | 28.5 | 0.38 (0.18 to 0.58) | <0.001 (<0.001) |

Prevalence classes are defined on Day 85 samples. The five most abundant species are P. copri, F. prausnitzii, P. stercorea, Bacteroides and Bifidobacterium.

**Table S8.** Consequence arm (exploratory): richness and log *P. stercorea* by illness burden and recency.

| Group | n | Species richness | log (count + 1) P. stercorea |
|---|---|---|---|
| Infections to day 85: 0 | 157 | 101.5 | 5.87 |
| Infections to day 85: 1 | 180 | 95.0 | 5.44 |
| Infections to day 85: 2 | 75 | 91.4 | 4.99 |
| Infections to day 85: 3+ | 40 | 95.1 | 5.08 |
| Last episode ended: ongoing/at sample | 26 | 88.7 | 4.39 |
| Last episode ended: 1-14 d | 48 | 89.0 | 5.00 |
| Last episode ended: 15-42 d | 127 | 96.8 | 5.22 |
| Last episode ended: >42 d | 94 | 94.6 | 5.75 |

Adjusted for age, sex, height-for-age, arm, season and location: richness changed by −1.1 species per additional infection (95% CI −3.3 to 1.2; p = 0.35), by −0.13 per illness day (p = 0.49) and by +0.09 per day since the last episode ended (p = 0.12).

**Table S9.** *P. stercorea* and ARI frequency in earlier analyses of this cohort (negative binomial; Day 85) [4,5].

| Model | IRR (95% CI) | Detail |
|---|---|---|
| P. stercorea (log), no richness adjustment (n = 447) | 0.946 (0.913–0.981) | beta = −0.0556, p = 0.002 |
| P. stercorea (log), richness-adjusted (n = 447) | 0.942 (0.907–0.979) | beta = −0.0594, p = 0.002 |
| P. stercorea (log), joint with P. copri (n = 447) | 0.951 (0.916–0.987) | p = 0.008; r with P. copri = 0.32 |

| Model | IRR (95% CI) | Detail |
|---|---|---|
| P. copri (log), joint model | 0.972 (0.913–1.038) | Not significant (p = 0.381) |

**Table S10.** Similarity matrix summary (see also Fig. 4a,b).

| Item | Value |
|---|---|
| Species columns | 488 (479 in matrix: 259 species-level, 220 genus-level; 9 not mapped) |
| Genomes used | 4,222 of 26,868 |
| Mean similarity, same genus / different genus | 0.80 / 0.38 |
| Share of counts covered per sample | median about 100% |
| Spearman ρ, richness vs redundancy at q = 0 / 1 / 2 | 0.93 / 0.63 / 0.33 |

**Table S11.** Spaceflight context: findings of the ISS astronaut study [6] and the Twins Study [7], as read from the source papers.

| Item | Finding |
|---|---|
| Design (ISS) | Nine astronauts, six to twelve months on the ISS; stool in flight from five (four opted out); 16S V4 sequencing; ten sampling times before, during and after flight. |
| Gut diversity | Shannon diversity and richness increased significantly in flight and returned to baseline after return. The exception was AstB, who had the highest pre- and post-flight diversity and richness and no significant change; four of five stool-sampled astronauts showed an increase. |
| Composition | Gut composition became more similar across four astronauts in flight; a few genera (Pseudobutyrivibrio, Dorea, Ruminococcus 2, Bifidobacterium, Blautia, Fusicatenibacter, Akkermansia) accounted for more than 90% of the reduction in variation. |
| Taxa | 17 gut genera changed (FDR < 0.05), 13 of them Firmicutes; more than five-fold fall in Akkermansia and Ruminococcus, about three-fold in Pseudobutyrivibrio and Fusicatenibacter; most reverted after return, except two genera. |
| Immune markers | By flight day 180, MCP-1, IL-8, IL-1b and MIP-1b increased, with IL-2 and IL-1ra; levels returned within two months. A Fusicatenibacter OTU correlated negatively with IL-8, IL-1b and TNF-alpha. VZV reactivation was detected in saliva of four crew members. |
| Recovery | Gut composition became similar to pre-flight within two months of return; skin and nose shifts persisted at least 60 days. |
| Authors' caveats | Exploratory; nine participants (five for stool); shared ISS diet a plausible driver of convergence; higher baseline diversity in AstB "possibly" conferring resilience, not tested. |
| Data access | Per-sample diversity is shown only in figures (Fig. 3 of the paper and its supplementary figures); no numeric table is published. Raw 16S reads are held in the NASA Life Sciences Data Archive by data request, because they are human-subject data. |
| Twins Study | Gut diversity unchanged; Firmicutes:Bacteroidetes ratio shifted in flight and returned to baseline. |

**Table S12.** Independent re-derivation of the Day 85 richness–infection association from the R1-only ASV table (separate vsearch pipeline: first 10,000 R1 reads per sample, UNOISE denoising, 99% identity mapping, no taxonomy). Richness is the expected number of ASVs in a random subsample of the stated number of reads (analytic rarefaction).

| Reads | Children (never / ill) | Mean ASV richness (never / ill) | Cohen's d | Welch p | Odds ratio of infection per SD (95% CI; p) | Post-sampling rate ratio per SD (95% CI; p) |
|---|---|---|---|---|---|---|
| 3000 | 125 / 303 | 324.2 / 290.2 | 0.37 | <0.001 | 0.76 (0.59–0.98; 0.037) | 0.69 (0.50–0.94; 0.018) |

| Reads | Children (never / ill) | Mean ASV richness (never / ill) | Cohen's d | Welch p | Odds ratio of infection per SD (95% CI; p) | Post-sampling rate ratio per SD (95% CI; p) |
|---|---|---|---|---|---|---|
| 5000 | 125 / 303 | 407.8 / 362.2 | 0.39 | <0.001 | 0.75 (0.58–0.97; 0.028) | 0.68 (0.50–0.94; 0.018) |
| 7000 | 110 / 253 | 462.9 / 409.3 | 0.39 | <0.001 | 0.71 (0.53–0.94; 0.016) | 0.66 (0.47–0.92; 0.016) |

ASV table: 9,038 ASVs in 442 Day 85 samples, 9 with no mapped reads; children are those with a sample at or above the stated depth and a Day 85 classification. Models adjust for age, sex, height-for-age, arm, season and location; the post-sampling model also adjusts for infections to day 85 and any episode within 14 days before the sample. ASV richness correlated with species-level richness (Spearman rho 0.90, n = 428). Same children and samples as the main analysis: a check on assignment and pipeline, not an independent cohort.

### Supplementary figures

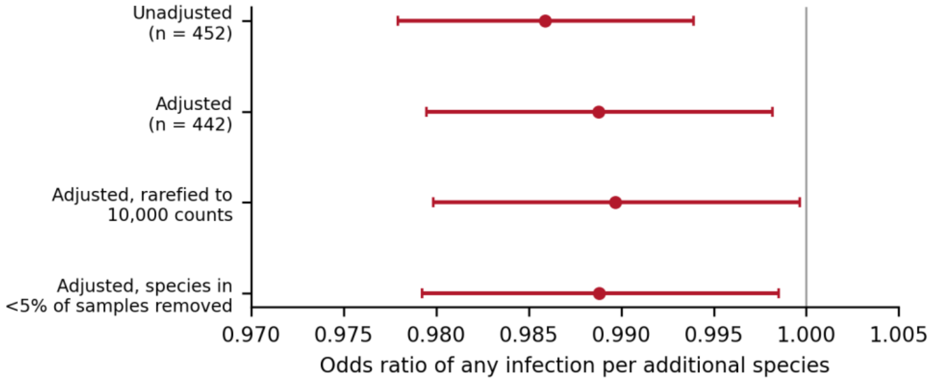


**Figure S1.** Robustness of the richness–illness association at Day 85. Odds ratios of any infection per additional species (95% CI): unadjusted; adjusted; adjusted with richness rarefied to 10,000 counts; adjusted after removing species present in under 5% of samples.

### References

1. Douglas GM, Maffei VJ, Zaneveld JR, et al. PICRUSt2 for prediction of metagenome functions. Nat Biotechnol 2020;38:685–8.

2. Parks DH, Chuvochina M, Rinke C, et al. GTDB: an ongoing census of bacterial and archaeal diversity through a phylogenetically consistent, rank normalized and complete genome-based taxonomy. Nucleic Acids Res 2022;50:D785–94.

3. Leinster T, Cobbold CA. Measuring diversity: the importance of species similarity. Ecology 2012;93:477–89.

4. Ofordile O, Pereira DIA, Prentice AM, Wagner J. Gut Prevotella stercorea associates with protection against infection in rural African children. Nat Commun 2025;16:11101.

5. Ofordile O. Prevotella stercorea links gut microbiome ecology to respiratory infection protection through host-context-dependent, richness-independent associations. BMC Med Genomics 2026.

6. Voorhies AA, Ott CM, Mehta S, et al. Study of the impact of long-duration space missions at the International Space Station on the astronaut microbiome. Sci Rep 2019;9:9911.

7. Garrett-Bakelman FE, Darshi M, Green SJ, et al. The NASA Twins Study: a multidimensional analysis of a year-long human spaceflight. Science 2019;364:eaau8650.
